# Integrative metabolomics differentiate coronary artery disease, peripheral artery disease, and venous thromboembolism risks

**DOI:** 10.1101/2023.06.21.23291103

**Authors:** Jiwoo Lee, Thomas Gilliland, Satoshi Koyama, Tetsushi Nakao, Jacqueline Dron, Kim Lannery, Megan Wong, Gina M. Peloso, Whitney Hornsby, Pradeep Natarajan

## Abstract

**Rationale:** Arterial and venous cardiovascular conditions, such as coronary artery disease (CAD), peripheral artery disease (PAD), and venous thromboembolism (VTE), are genetically correlated. Interrogating distinct and overlapping mechanisms may shed new light on disease mechanisms.

**Objective:** In this study, we aimed to: identify and compare (1) epidemiologic and (2) causal, genetic relationships between metabolites and CAD, PAD, and VTE.

**Methods:** We used metabolomic data from 95,402 individuals in the UK Biobank, excluding individuals with prevalent cardiovascular disease. Logistic regression models adjusted for age, sex, genotyping array, first five principal components of ancestry, and statin use estimated the epidemiologic associations of 249 metabolites with incident CAD, PAD, or VTE. Bidirectional two-sample Mendelian randomization (MR) estimated the causal effects between metabolites and cardiovascular phenotypes using genome-wide association summary statistics for metabolites (N = 118,466 from UK Biobank), CAD (N = 184,305 from CARDIoGRAMplusC4D 2015), PAD (N = 243,060 from Million Veterans Project) and VTE (N = 650,119 from Million Veterans Project). Multivariable MR (MVMR) was performed in subsequent analyses.

**Results:** We found that 194, 111, and 69 metabolites were epidemiologically associated (P < 0.001) with CAD, PAD, and VTE, respectively. Metabolomic profiles exhibited variable similarity between disease pairs: CAD and PAD (N = 100 shared associations, R^2^ = 0.499), CAD and VTE (N = 68, R^2^ = 0.455), and PAD and VTE (N = 54, R^2^ = 0.752). MR revealed 28 metabolites that increased risk for both CAD and PAD and 2 metabolites that increased risk for CAD but decreased risk for VTE. Despite strong epidemiologic overlap, no metabolites had a shared genetic relationship between PAD and VTE. MVMR revealed several metabolites with shared causal effects on CAD and PAD related to cholesterol content within very-low-density lipoprotein particles.

**Conclusions:** While common arterial and venous conditions are associated with overlapping metabolomic profiles, MR prioritized the role of remnant cholesterol in arterial diseases but not venous thrombosis.

## Introduction

Cardiovascular diseases, including coronary artery disease (CAD), peripheral artery disease (PAD), and venous thromboembolism (VTE), remain the leading causes of morbidity and mortality worldwide.^1^ Studies suggest mechanistic overlap between CAD and PAD with atherosclerosis as a principal driver of both.^2,3^ In the past decade, genome-wide association studies (GWAS) of CAD, PAD, and VTE have probed the genetic architectures of and between these three phenotypes.^2,4–6^ In addition, CAD, PAD, and VTE often co-occur – upwards of 42% of individuals with CAD also have PAD.^7^ Understanding shared and distinct pathways may facilitate therapeutic interventions toward key aspects of disease mechanisms.

Assessment of the human metabolome is an attractive approach to study mechanistic etiologies, as metabolomics capture exposures from several sources, including genetic, dietary, and lifestyle profiles, that all influence an individual’s lifetime risk of various cardiovascular diseases.^8^ Identifying upstream metabolites that affect an individual’s risk of CAD, PAD, or VTE and vice versa (i.e., identifying metabolites that are affected by having CAD, PAD, or VTE) are critical to better understand mechanistic etiology. Recent GWAS performed on large-scale metabolomics data^9^ may help to probe the role of the metabolome in explaining the genetic overlap between CAD, PAD, and VTE.^9^ Examination of genetic pleiotropy offers a new opportunity to shed new light on disease pathogenesis.^10^

Mendelian randomization (MR) can estimate causal effects between metabolites and CAD, PAD, and VTE. MR is a causal inference approach that uses genetic instruments as instrumental variables to prioritize potentially causal relationships between exposures and outcomes, with corresponding lowering risks of confounding and reverse causation.^11^ MR is particularly well-suited for estimating the causal effects of metabolites, especially given their strong heritability^12^ and recent in-depth molecular profiling of metabolites in the UK Biobank.^13,14^ Given widespread correlation of metabolites, we leverage methodologic advances accounting for horizontal pleiotropic effects and potential reverse causation.

In this study, we aimed to: (1) identify and compare associations of metabolites with CAD, PAD, and VTE and (2) identify and compare causal effects of metabolites with CAD, PAD, and VTE. Specifically, we performed association analyses and bidirectional two-sample MR analyses in the UK Biobank with 95,402 individuals with in-depth molecular profiling of metabolites and non-overlapping genetic data for the cardiovascular outcomes from large-scale GWAS consortia.

## Methods

### Study cohort

The UK Biobank study cohort has been described previously.^14^ Briefly, the UK Biobank is a large population-based prospective study that contains genotype and phenotype data from 502,639 individuals living in the United Kingdom and recruited between 2006 and 2010.^14^ Individuals who were closely related, withdrew consent, or had prevalent CAD, PAD, or VTE at baseline were excluded. CAD, PAD, and VTE were defined by the International Classification of Diseases (ICD) system, specifically ICD-10 codes (**Supplementary Table 1**). In CARDIoGRAMplusC4D 2015, CAD cases were similarly defined by an inclusive CAD diagnosis (e.g., myocardial infarction, acute coronary syndrome, chronic stable angina, or coronary stenosis > 50%).^15^ In the Million Veterans Program, PAD and VTE were similarly defined by ICD-9, ICD-10, and Current Procedural Terminology (CPT) codes (**Supplementary Table 1**).^2,6^

A random subset of non-fasting baseline plasma samples from 118,466 individuals and 1,298 repeat visit samples were measured for 249 metabolomic biomarkers using a high-throughput nuclear magnetic resonance (NMR) metabolomic biomarker profiling platform developed by Nightingale Health (Helsinki, Finland) in the UK Biobank.^9^ The metabolites span multiple metabolic pathways, including lipoprotein lipids in 14 subclasses, fatty acids and fatty acid compositions, as well as various low-molecular weight metabolites, such as amino acids, ketone bodies, and glycolysis metabolites, all quantified in molar concentration units.^9^ A total of 95,402 individuals were included in this study.

### Genetic correlation between cardiovascular phenotypes

After variant filtering (e.g., INFO > 0.9 and minor allele frequency (MAF) > 0.01), linkage disequilibrium score regression (LDSC) with default parameters were used to calculate the genetic correlation between CAD and PAD, CAD and VTE, and PAD and VTE using HapMap3 single nucleotide polymorphisms (SNPs) in the 1000 Genomes Project European reference panel.^16^ Summary statistics for CAD were obtained from CARDIoGRAMplusC4D 2015 that filtered out individuals from the UK Biobank,^15^ and summary statistics for PAD and VTE were obtained from the Million Veterans Program.^2,6^

### Association analysis of metabolites with cardiovascular phenotypes

To estimate epidemiologic associations of 249 metabolites with CAD, PAD, and VTE, we constructed logistic models regressing metabolite measurements on CAD, PAD, and VTE coded as a binary indicator for incident event, with covariates including age, sex, genotyping array, five genetic principal components, and self-reported statin use. A correction factor of 0.5 multiplied by the minimum non-zero value of each metabolite was applied prior to log transformation of metabolite values. Transformed metabolite values were scaled with a mean of 0 and a standard deviation of 1. Effect sizes and standard errors were reported, and alpha was defined as 0.05 / 41 = 0.001. Bonferroni correction for multiple-hypothesis testing is too strict given the high degree of correlation of the 249 metabolites, as previously described.^17^ Consistent with prior studies to minimize both type I and type II errors, we adjusted for the number of principal components (N = 41) that explain 99% of the variance in the data (e.g., 0.05 / 41 = 0.001).^17^ To estimate the correlations of associations between CAD, PAD, and VTE, we calculated the Pearson correlation coefficients of z-scores from logistic regression as a measure of metabolomic profile overlap.

### Mendelian randomization of metabolites and cardiovascular phenotypes

Mendelian randomization (MR) is a method that uses genetic variants as instrumental variables to prioritize causal relationships between exposures and outcomes.^11^ MR relies on several assumptions: (1) genetic instruments must be robustly associated with the exposure, (2) genetic instruments must not be associated with confounders, and (3) genetic instruments must not influence the outcome except through the exposure.^11^ Two-sample MR utilizes GWAS summary statistics in non-overlapping cohorts for exposures and outcomes.^18^ We used two non-overlapping cohorts for metabolites and cardiovascular outcomes. Summary statistics for metabolites were obtained from publicly available GWAS summary statistics from the OpenGWAS platform (https://gwas.mrcieu.ac.uk/), which contained 249 metabolites from the UK Biobank measured by Nightingale Health (**Supplementary Table 2**).^19^ Summary statistics were not adjusted for statin use. Summary statistics for CAD were obtained from CARDIoGRAMplusC4D 2015,^15^ and summary statistics for PAD and VTE were obtained from the Million Veteran Program.^2,6^

We performed bidirectional, two-sample MR using the TwoSampleMR^20,21^ R package to estimate the causal effects between metabolites and CAD, PAD, and VTE. All analyses were consistent with current recommendations for MR.^22^ We performed linkage disequilibrium (LD) clumping with a window > 10000 kb and an R^2^ > 0.001 Genetic instruments were defined to be genetic variants that were significant at the genome-wide significance level after LD clumping (P < 5×10^-8^). A median of 66 SNPs were used as the instrumental variables for the metabolites (SD = 19.9, minimum N SNPs = 7, maximum N SNPs = 112), and instrumental variables were robust, with all F-statistics greater than 50. We utilized the inverse variance weighted method, which regresses associations between genetic instruments and outcomes upon associations between genetic instruments and exposures.^23^

In subsequent sensitivity analyses, we utilized the MR Egger, weighted median, simple mode, and weighted mode methods as well, as such methods may be more robust to pleiotropy. We also conducted tests for heterogeneity and horizontal pleiotropy. In MR, a test for heterogeneity assesses the compatibility of instrumental variable estimates based on individual genetic variants and is calculated using Cochran’s Q test on the causal estimates from individual genetic variants.^24^ We considered P > 0.05 to indicate no statistical evidence of heterogeneity. A test for horizontal pleiotropy is calculated using the MR Egger regression method, as the slope coefficient estimates the causal effect that is consistently asymptotically even if individual genetic variants have pleiotropic effects.^24^ We considered P > 0.05 to indicate no statistical evidence of horizontal pleiotropy.

In the first analysis, in which metabolites were exposures and CAD, PAD, and VTE were outcomes, genetic instruments were clumped and selected based on the above criteria (e.g., P < 5×10^-8^ for metabolite association from GWAS). In the second analysis, in which metabolites were outcomes and CAD, PAD, and VTE were exposures, genetic instruments were clumped and selected based on the above criteria (e.g., P < 5×10^-8^ for CAD, PAD, or VTE association from GWAS). To estimate the correlations of causal effects between CAD, PAD, and VTE, we calculated the Pearson correlation coefficients of betas from MR as a measure of metabolomic profile overlap.

Given the high degree of correlation among metabolites, we also performed multivariable Mendelian randomization (MVMR), which uses genetic variants for two or more exposures to simultaneously estimate the causal effect of each exposure on the outcome, controlling for the effect of other included exposures. MVMR requires the same assumptions as univariable MR, but the genetic instruments must be associated with the set of exposures rather than the single exposure, but it is not necessary for each genetic instrument to be significantly associated with every exposure.^25^ In other words, instruments are selected for each exposure and then all exposures for such instruments are regressed against the outcome together, weighting for the inverse variance of the outcome. MVMR was performed using the set of metabolites that were identified to have causal effects in the first analysis, such that the exposures included the set of significant metabolites and the outcomes included CAD, PAD, and VTE separately, in order to identify metabolites that had both strong and independent causal effects.

All analyses were performed using R version 4.1 and all plots were generated using the R package “ggplot2”. A heatmap was generated using the R package “gplots”.

## Results

We estimated associations of 249 metabolites with CAD, PAD, and VTE in 95,402 individuals in the UK Biobank, which included 3,209 CAD cases, 846 PAD cases, and 1,474 VTE cases (**Figure 1**). The study cohort of 95,402 comprised of 233,202 (55%) females with a mean (standard deviation) age of 56.3 (8.0) years, and 57,948 (14%) individuals prescribed statins (**Table 1**). The GWAS summary statistics for CAD included 60,801 cases and 123,504 controls across 48 non-overlapping studies;^15^ for PAD included 31,307 cases and 211,753 controls from the Million Veterans Program;^2^ and for VTE included 26,066 cases and 624,053 controls from the Million Veterans Program.^6^ We calculated the genetic correlations across the three sets of summary statistics. CAD and PAD shared the highest genetic correlation (Rg = 0.6615, SE = 0.044, P = 4.51×10^-51^), followed by PAD and VTE (Rg = 0.2463, SE = 0.053, P = 3.28×10^-6^) and CAD and VTE (Rg = 0.1300, SE = 0.0480, P = 6.80×10^-3^).

**Figure 1.**
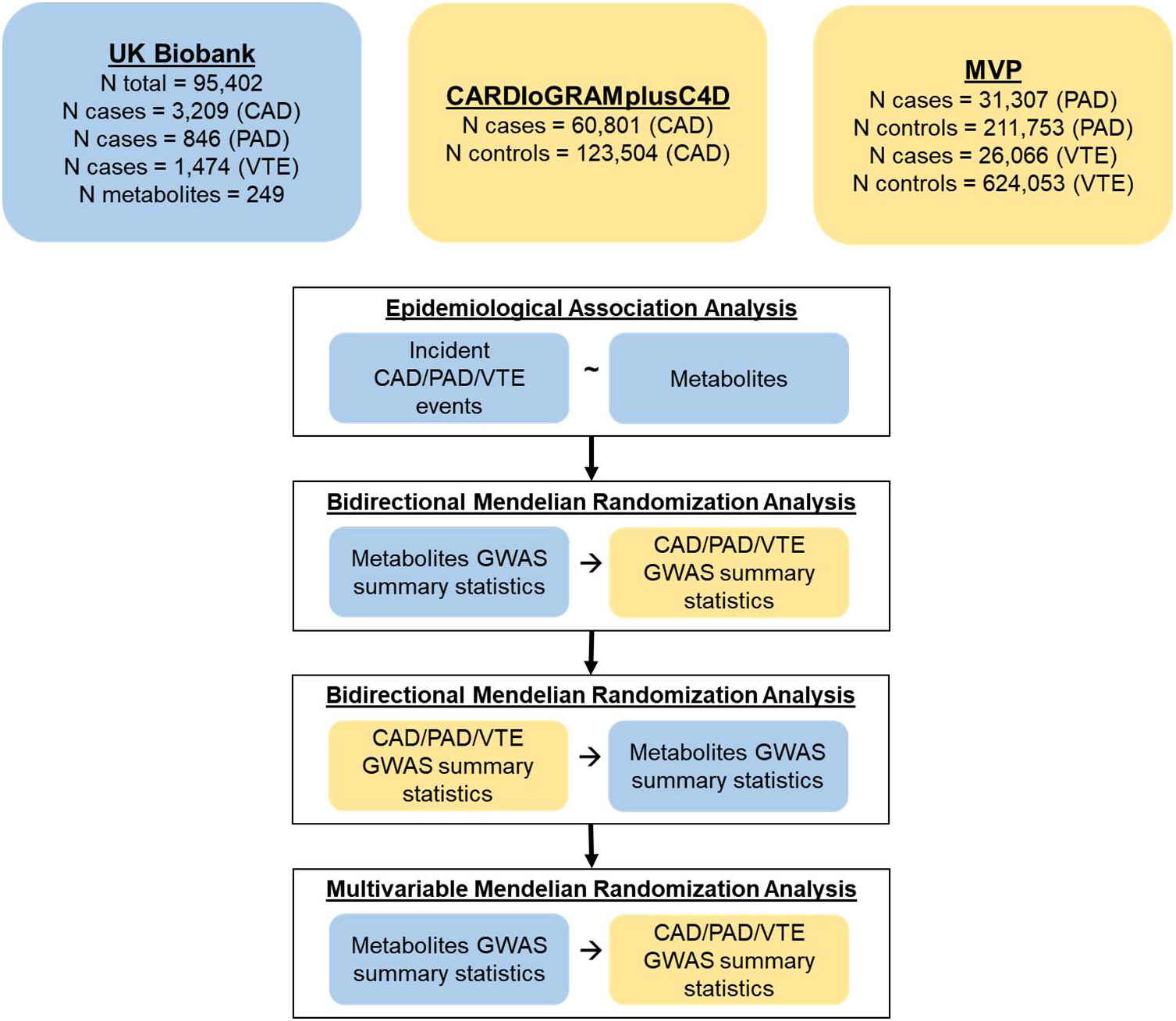
Study schematic. (A) Epidemiological association analysis was performed using individual-level data from the UK Biobank, regressing incident coronary artery disease (CAD)/peripheral artery disease (PAD)/venous thromboembolism (VTE) events on metabolite levels. (B) Bidirectional Mendelian randomization analysis was performed using metabolite summary statistics from the UK Biobank as exposures and CAD/PAD/VTE summary statistics from either CARDIoGRAMplusC4D 2015 or Million Veterans Program (MVP) as outcomes, and (C) vice versa. (D) Taking only the metabolite with significant causal effects from (B), multivariable Mendelian randomization analysis was performed using metabolite summary statistics from the UK Biobank as exposures and CAD/PAD/VTE summary statistics from either CARDIoGRAMplusC4D 2015 or MVP as outcomes.

**Table 1.** Baseline characteristics of analyzed UK Biobank participants. Individuals with prevalent CAD, PAD, or VTE were excluded, leaving 3,209, 846, 1,474 incident coronary artery disease (CAD), peripheral artery disease (PAD), and venous thromboembolism (VTE) cases. The baseline characteristics are reported below.

|  |  |
| --- | --- |
| **N** | 95,402 |
| **Age (SD)** | 56.3 (8.0) |
| **N Female (%)** | 52,065 (54.6%) |
| **N European (%)** | 89,847 (94.2%) |
| **N Hypertension (%)** | 26,507 (27.8%) |
| **N Diabetes (%)** | 1,974 (2.1%) |
| **N Current Smoker (%)** | 9,754 (10.2%) |
| **N Statin Users** | 13,204 (13.8%) |
| **Total cholesterol, mg/dL (SD)** | 221.5 (43.4) |
| **Low-density lipoprotein cholesterol, mg/dL (SD)** | 138.7 (33.2) |
| **High-density lipoprotein cholesterol, mg/dL (SD)** | 56.3 (14.7) |
| **Triglycerides, mg/dL (SD)** | 153.2 (89.1) |
| **N Incident CAD (%)** | 3,209 (3.36%) |
| **N Incident PAD (%)** | 846 (0.887%) |
| **N Incident VTE (%)** | 1,474 (1.55%) |

### Epidemiologic associations reveal overlap of metabolites between cardiovascular phenotypes

We found 194, 111, and 69 metabolites epidemiologically associated with CAD, PAD, and VTE, respectively (**Figure 2, Supplementary Table 3**). Phospholipids / total lipids in large VLDL (odds ratio (OR) = 1.62, 95% CI = [1.39, 1.89], P = 5.04×10^-10^) and apolipoprotein B / apolipoprotein A1 (OR = 1.43, 95% CI = [1.37, 1.49], PS = 7.29×10^-72^) were among the top metabolites associated with increased CAD risk, while cholesteryl esters in high-density lipoprotein (HDL) were associated with decreased CAD risk (OR = 0.73, 95% CI = [0.70, 0.75], P = 1.82×10^-58^). Glycoprotein acetyls were associated with increased PAD risk (OR = 1.40, 95% CI = [1.31, 1.50], P = 7.57×10^-24^) and degree of unsaturation of all fatty acids was associated with decreased PAD risk (OR = 0.72, 95% CI = [0.68, 0.77], P = 1.47×10^-22^). Phospholipids / total lipids in medium HDL was associated with increased VTE risk (OR = 1.21, 95% CI = [1.14, 1.29], P = 2.29×10^-9^) while albumin was associated with decreased VTE risk (OR = 0.80, 95% CI = [0.77 0.84], P = 1.11×10^-18^).

**Figure 2.**
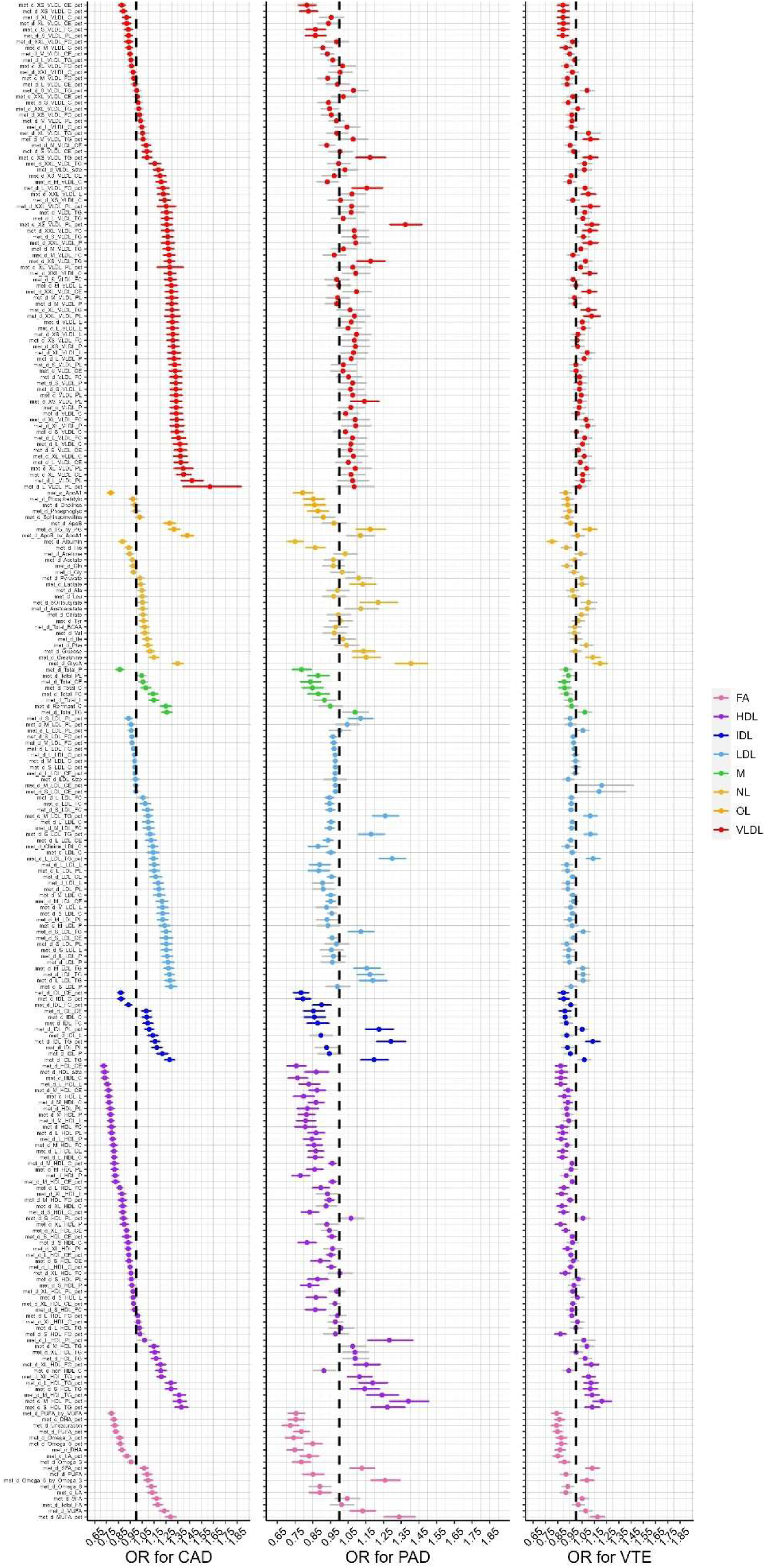
Epidemiological associations between metabolites and incident cardiovascular outcomes. Associations were estimated using logistic regression on 95,402 individuals in the UK Biobank and 7 classes of metabolites: VLDL = very low-density lipoprotein, FA = fatty acid, HDL = high-density lipoprotein, IDL = intermediate-density lipoprotein, LDL = low-density lipoprotein, M = miscellaneous, NL = non-lipid metabolite, and OL = other lipid metabolite. Bars oriented upwards indicate a positive association while those oriented inwards indicate a negative association. Error bars represent 95% confidence intervals. Transparent bars represent associations that were not significant based on a multiple testing correction of P < 0.05 / 41. Coronary artery disease (CAD) was associated with 194 metabolites. Peripheral artery disease (PAD) was associated with 111 metabolites. Venous thromboembolism (VTE) was associated with 69 metabolites.

Next, we estimated the overlap of metabolomic associations across CAD, PAD, and VTE. There were 52 metabolites that had significant (P < 0.001) associations with all three cardiovascular phenotypes, of which 50 metabolites had the same direction of effect and 2 metabolites had the opposite direction of effect of CAD compared to PAD and VTE. CAD and PAD shared 48 significant metabolites (30 concordant, 18 discordant), CAD and VTE shared 16 metabolites (16 concordant, 0 discordant), and PAD and VTE shared 2 metabolites (2 concordant, 0 discordant) (**Figure 3**). Epidemiologic metabolomic profiles were most similar between PAD and VTE (R^2^ = 0.75, P = 8.43×10^-775^), followed by CAD and PAD (R^2^ = 0.50, P = 5.71×10^-395^) and CAD and VTE (R^2^ = 0.46, P = 2.14×10^-345^). In other words, while CAD and PAD shared the highest genetic correlation, PAD and VTE shared the highest similarity of metabolomic associations (**Supplementary Figure 1**).

**Figure 3.**
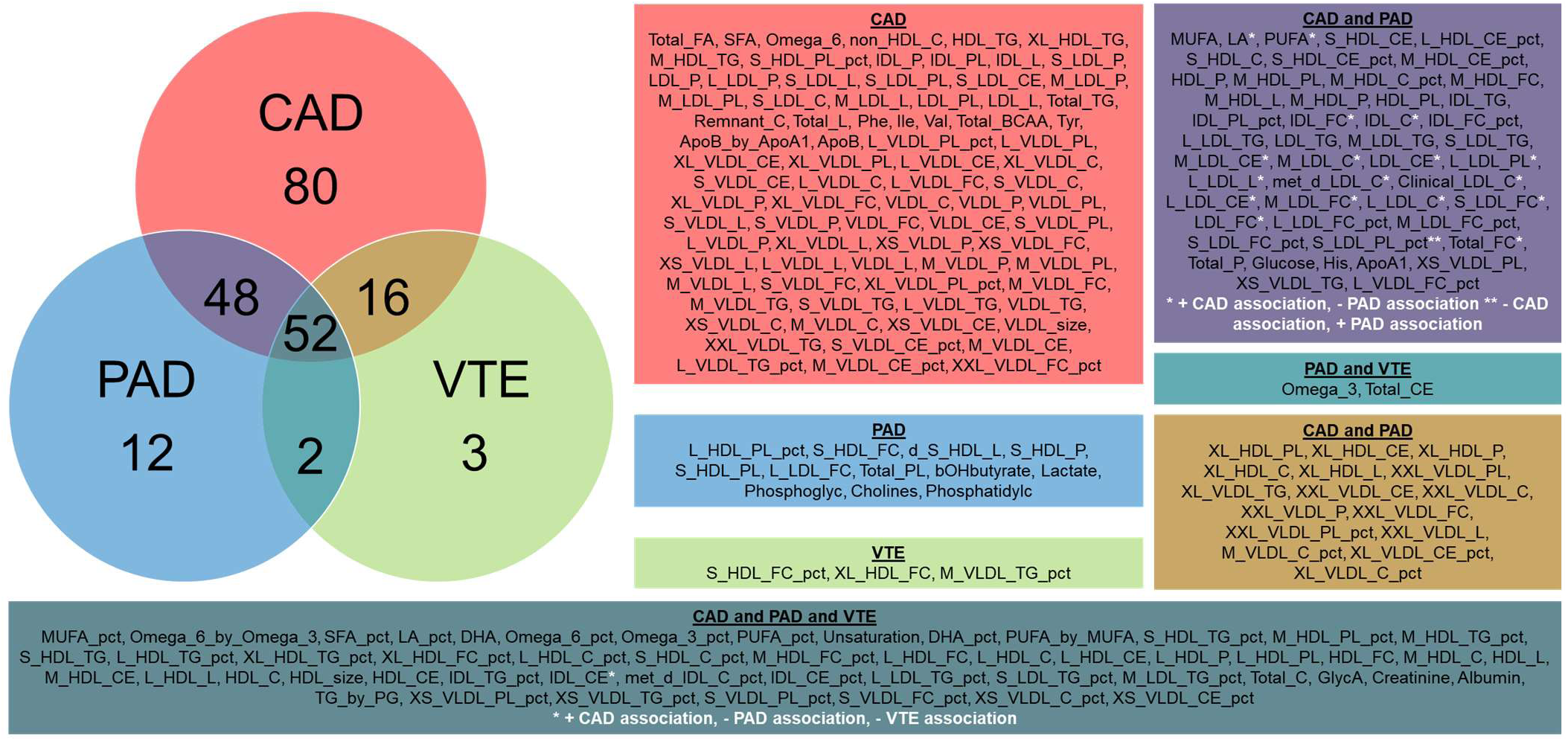
Venn diagram of metabolites with shared significant associations between coronary artery disease (CAD), peripheral artery disease (PAD), and venous thromboembolism (VTE) estimated using logistic regression. Associations were estimated using logistic regression on 95,402 individuals in the UK Biobank. Metabolites that were associated with more than one cardiovascular phenotype but demonstrated an opposite direction of effect were indicated with white asterisks.

### Estimation of causal effects of metabolites on cardiovascular phenotypes reveal overlap between coronary artery disease and peripheral artery disease

Then, we used bidirectional MR to identify potential causal relationships between metabolites and CAD, PAD, and VTE (**Figure 4, Supplementary Figure 2**). We found 82 metabolites that increased and 5 that decreased risk for CAD, 25 metabolites that increased and 11 that decreased risk for PAD, and 2 metabolites that decreased risk for VTE (P < 0.001). Many of the HDL traits were found to have causal effects on PAD while many of the IDL traits, and larger-sized lipid traits, were found to have causal effects on CAD. Most VLDL and LDL traits were shared by both CAD and PAD. We also performed sensitivity analyses using the MR Egger, weighted median, simple mode, and weighted mode methods (**Supplementary Tables 4-9**), as well was testing for heterogeneity (**Supplementary Table 10**) and horizontal pleiotropy (**Supplementary Tables 11**) to assess the robustness of our results and instrumental variables. Most metabolites showed no statistical evidence (P > 0.05) for horizontal pleiotropy in their causal effects on CAD (N = 175 metabolites without horizontal pleiotropy), PAD (N = 200), and VTE (N = 246).

**Figure 4.**
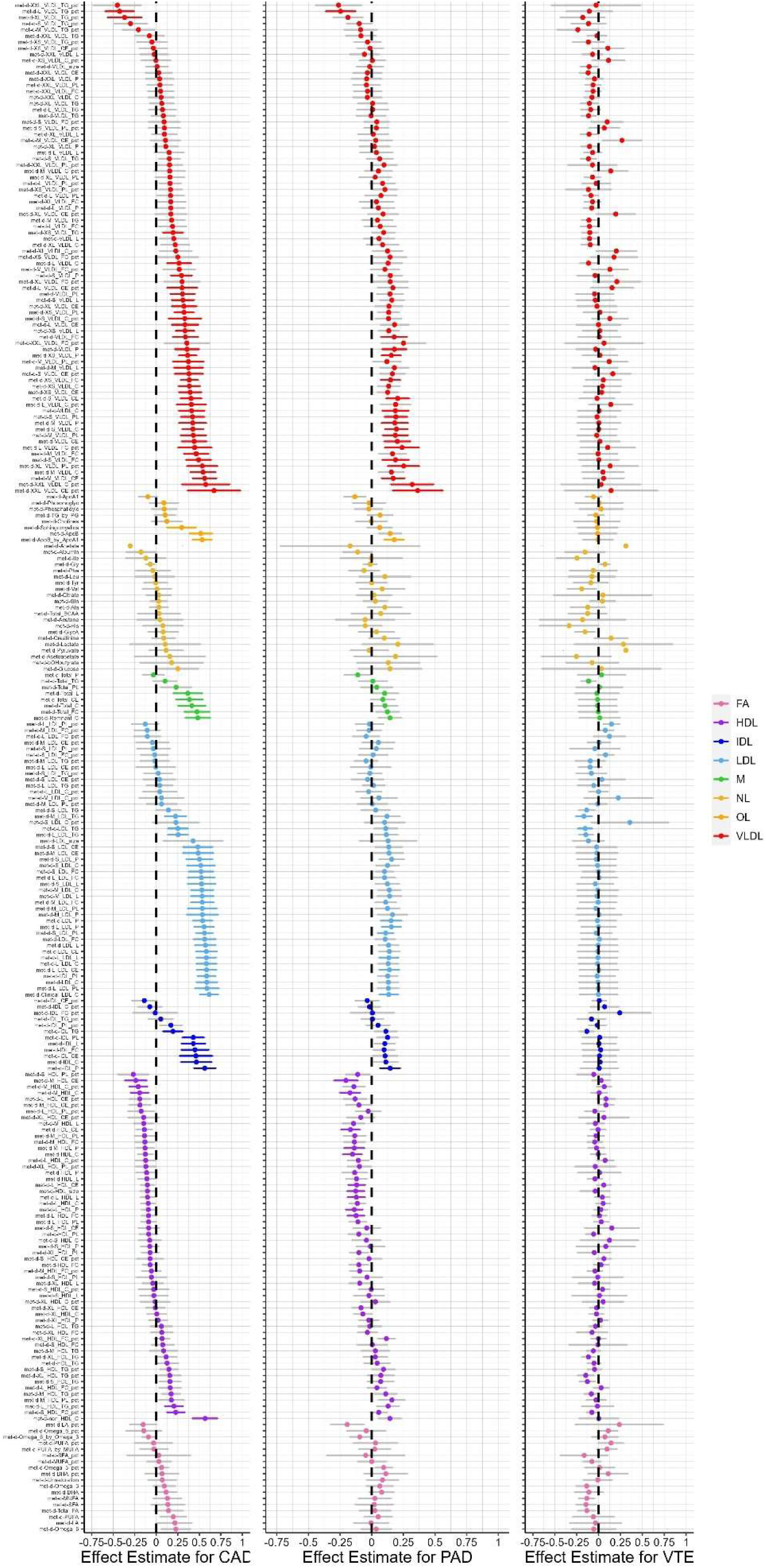
Forest plots of estimated causal effects of genetically-determined metabolites on cardiovascular outcomes estimated using bidirectional Mendelian randomization. Bidirectional, two-sample inverse variance weighted Mendelian randomization using summary statistics from the UK Biobank and CARDIoGRAMplusC4D 2015 or Million Veterans Program (MVP) was performed to estimate causal effects of metabolites on coronary artery disease (CAD), peripheral artery disease (PAD), and venous thromboembolism (VTE). Bars are effect estimates from Mendelian randomization. Error bars represent 95% confidence intervals. Transparent bars represent associations that were not significant based on a multiple testing correction of P < 0.05 / 41. There were 87, 36, and 2 metabolites with a causal effect on CAD, PAD, and VTE, respectively.

Conversely, while we found that CAD increased 14 metabolites and decreased 6 metabolites, we found no metabolites affected by PAD or VTE. Interestingly, 5 of the 14 metabolites had bidirectional, increased causality with CAD (e.g., the metabolite increased risk of CAD and CAD also increased levels of the metabolite) and were all involved in cholesterol concentrations in large VLDL and extremely large VLDL (**Supplementary Figure 3**). In addition, 2 of the 6 metabolites had bidirectional, decreased causality with CAD (e.g., the metabolite decreased risk of CAD and CAD also decreased levels of the metabolite) and were all involved in triglyceride concentrations in large VLDL and very large VLDL (**Supplementary Figure 3**). For the 7 metabolites with bidirectional, causal effects with CAD, there were only one overlap in the variants driving the causal effect of the metabolite on CAD and the variants driving the causal effect of CAD on the metabolite: rs2519093 and rs7528419. The majority of genetic variants only had a causal effect on the metabolite or CAD in a single direction.

Causal metabolomic profiles were most similar between CAD and PAD (R^2^ = 0.43, P = 4.67×10^-156^), followed by PAD and VTE (R^2^ = 0.06, P = 3.55×10^-17^) and CAD and VTE (R^2^ = 0.03, P = 9.38×10^-11^). MR revealed 28 metabolites that increased risk for both CAD and PAD and 2 metabolites that increased risk for CAD but decreased risk for VTE (i.e., triglycerides in low-density lipoprotein (LDL), triglycerides in medium LDL). (**Figure 5**). There were no metabolites with shared causal relationships between PAD and VTE. Of the 28 metabolites shared between CAD and PAD, 64.3% (N = 18) were VLDL, 21.4% (N = 6) were LDL, 7.1% (N = 2) were HDL, 3.6% (N = 1) were IDL, and 3.6% (N = 1) were apolipoprotein traits. Many of the triglyceride-containing lipid traits that were epidemiologically associated with CAD and PAD did not maintain causal effects; rather, the 28 metabolites shared between CAD and PAD included many cholesterol-containing lipid traits but no triglyceride-containing lipid traits. Taken together with the epidemiologic association results, this suggests that while PAD and VTE maintain similar metabolomic associations, these effects are not robust under a causal framework such that CAD and PAD retain more similar causal metabolomic profiles (**Supplementary Figure 1**). Notably, many of the epidemiologic associations that were not maintained upon interrogation via MR included non-lipid traits, such as glucose and other amino acids.

**Figure 5.**
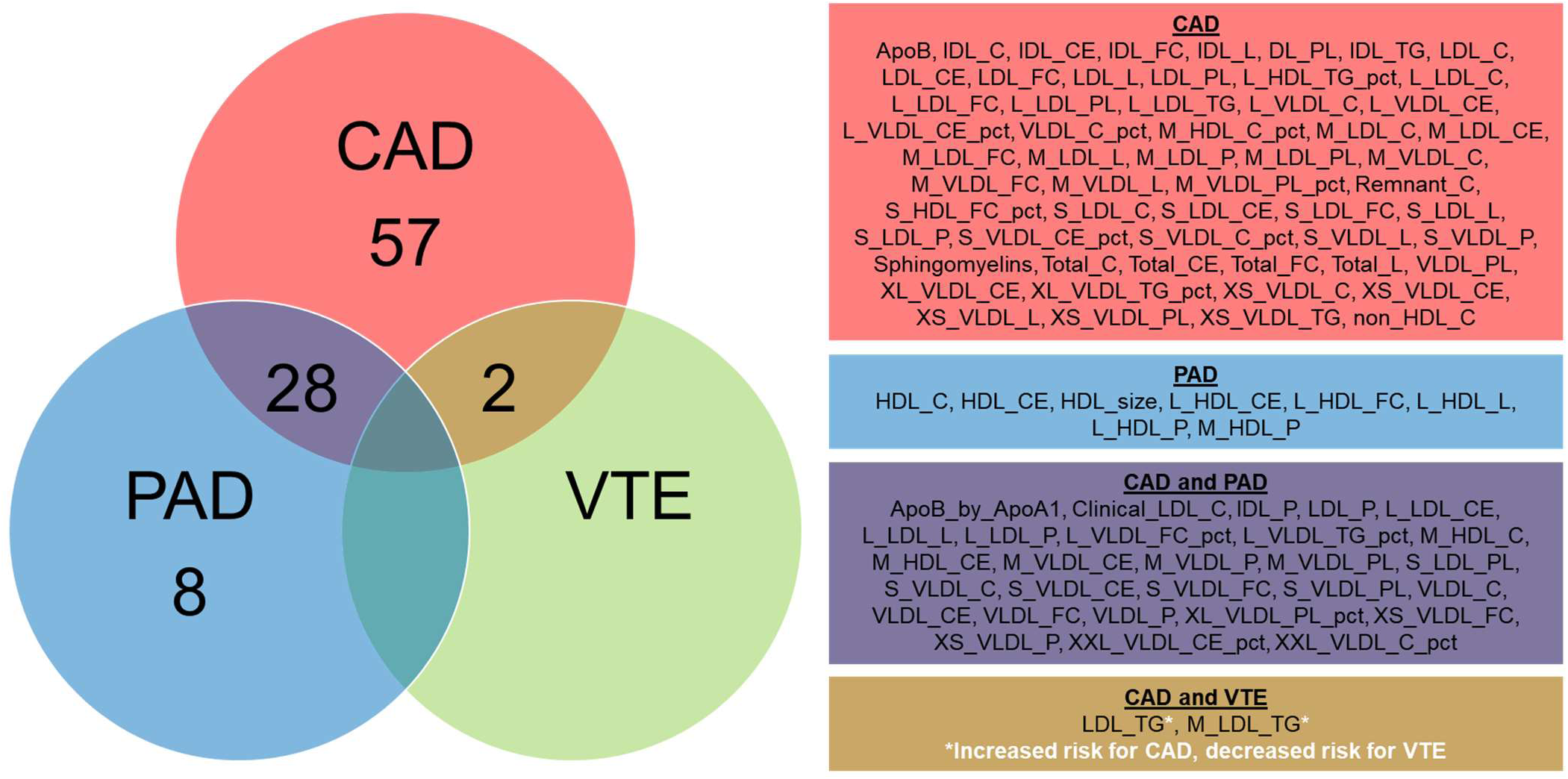
Venn diagram of metabolites with shared causal effects between coronary artery disease (CAD), peripheral artery disease (PAD), and venous thromboembolism (VTE) estimated using bidirectional Mendelian randomization. Bidirectional, two-sample inverse variance weighted Mendelian randomization using summary statistics from the UK Biobank and CARDIoGRAMplusC4D 2015 or Million Veterans Program (MVP) was performed to estimate causal effects of metabolites on CAD, PAD, and VTE. Metabolites that had causal effects on more than one cardiovascular phenotype but demonstrated an opposite direction of effect were indicated with white asterisks.

Given the high degree of correlation of metabolites among each other, we used MVMR to identify metabolites that had causal effects on CAD, PAD, and VTE independent of other metabolites (**Supplementary Tables 9-11**). In three models including the 87, 36, and 2 metabolites with causal effects on CAD, PAD, and VTE as exposures, we identified 7 metabolites with marginal (P < 0.05) independent causal effects on CAD (i.e., cholesteryl esters in VLDL, total lipids in small VLDL, concentration of medium VLDL particles, total lipids in lipoprotein particles, total lipids in LDL, triglycerides to total lipids ratio in large HDL, and total cholesterol minus HDL-C) and 5 on PAD (i.e., cholesteryl esters in small VLDL, phospholipids in small VLDL, free cholesterol in small VLDL, cholesterol in small VLDL, and apolipoprotein B / apolipoprotein A1). We did not identify any metabolites with independent causal effects on VTE. In a separate model excluding the 7 metabolites that had bidirectional, causal effects with CAD, we identified 8 metabolites with marginal (P < 0.05) independent causal effects on CAD (i.e., total lipids in LDL, cholesterol in IDL, cholesteryl esters in medium LDL, apolipoprotein B, free cholesterol in medium LDL, total lipids in lipoprotein particles, concentration of medium VLDL particles, and total lipids in IDL). There were three metabolites with marginal independent causal effects in both models including the primary model including all 87 metabolites and the secondary model excluding the 7 metabolites that had bidirectional, causal effects on CAD (i.e., total lipids in lipoprotein particles, total lipids in LDL, concentration of medium VLDL particles).

## Discussion

Here, we performed hypothesis-free association analyses and MR analyses to evaluate the metabolomic profiles of CAD, PAD, and VTE covering 249 metabolites in 95,402 individuals in the UK Biobank. This allowed us to compare metabolomic profiles between these three cardiovascular phenotypes to provide insight into potential similarities and differences in their metabolomic etiologies.

First, we found overlap in associated metabolomic profiles of CAD, PAD, and VTE via association analyses. We identified 194, 111, and 69 metabolites that were associated with CAD, PAD, and VTE, respectively, that were also consistent with well-established biology, such as the association between apolipoprotein B and apolipoprotein A1 with CAD. We compared the associated metabolites across CAD, PAD, and VTE and found that CAD and PAD shared 48 metabolites, CAD and VTE shared 16 metabolites, and PAD and VTE shared 2 metabolites - there were 52 metabolites shared by all three cardiovascular phenotypes. There was a high degree of similarity in metabolomic associations between PAD and VTE compared to CAD and VTE. Identification of epidemiologic associations provided a background comparison for subsequent estimation of causal effects.

Second, we found that overlap in the causal metabolic profiles of CAD and PAD, but not VTE, via MR analyses. This is consistent with the high co-occurrence of CAD and PAD, as well as the genetic correlation, reported by previous studies. ^2,4–7^ We identified several upstream metabolites that affected an individual’s risk of CAD (N = 87), PAD (N = 36), and VTE (N = 2). Specifically, 28 metabolites increased the risk for both CAD and PAD and 2 metabolites had opposite causal effects in CAD and VTE. Of the 28 metabolites many were VLDL traits, and indeed, VLDL pathways have been implicated in both CAD^26^ and PAD.^27^ VLDL is a precursor to LDL and is considered to be a marker of atherogenic lipoprotein remnants.^28^ Given that both CAD and PAD are characterized by the buildup of plaque in coronary or peripheral arteries, it is possible that VLDL, specifically the smaller VLDL particles that are closer in metabolomic identity to LDL, contributes to the development of atherosclerotic lesions in both arterial locations. The convergence of VLDL cholesterol concentrations between CAD and PAD metabolomic profiles strengthens this shared disease mechanism. This result supports the prioritization of atherogenic lipoprotein remnants as a high-yield therapeutic target for both CAD and PAD due to overlaps in lipid-related etiology. Furthermore, no triglyceride-containing lipid traits were shared between CAD and PAD, while many cholesterol-containing lipid traits were shared. Given that previous studies have suggested that triglycerides are not atherogenic, the overrepresentation of cholesterol-containing lipid traits further supports the role of lipid-related etiology underlying the atherosclerotic mechanisms of both CAD and PAD, specifically the cholesterol content of VLDL particles. ^29,30^

Thus, while the association analyses showed a high degree of metabolomic associations between PAD and VTE, causal effects of metabolites were shared between CAD and PAD to a greater degree than CAD and VTE. Lipid traits may not be as strongly involved in thrombus formation as it is in plaque formation, as VTE had few lipid traits with causal effects with CAD or PAD. Previous studies have highlighted carnitine species, glucose, phenylalanine, among other metabolites, but not lipid-related metabolites, as possibly related to VTE.^31^ We similarly found that several epidemiologic associations identified via association analyses, such as glucose and other amino acids, did not maintain robust causal effects upon interrogation via MR.

Third, we found that CAD may amplify the effects of some metabolites, particularly cholesterol and triglycerides within VLDL. Using bidirectional MR, we identified 20 downstream metabolites that had levels affected by CAD, but not PAD or VTE. Most of the 20 metabolites were involved in VLDL and LDL particles, and those that had positive bidirectional effects with CAD included cholesterol concentrations and those that had negative bidirectional effects with CAD included triglyceride concentrations. Regarding VTE, the null effects of metabolites on VTE and vice versa in a bidirectional MR analysis was previously reported for LDL, HDL, and TGs.^32^ Regarding CAD, this bidirectional effect may suggest that atherosclerosis itself may influence lipid metabolism. Tomas et al. found distinct metabolomic profiles in high-risk and stable atherosclerotic plaques consistent with different transcription levels of metabolic enzymes, suggesting that atherosclerosis itself may affect metabolite levels.^33^ Further functional studies are needed to evaluate this bidirectional effect.

The limitations of this study should be considered. First, the power and precision of MR is limited to the strength and availability of instrumental variables, such as for VTE. As more powerful GWAS are performed that produce more, high-quality instrumental variables, the MR results will necessarily become stronger and more robust. Nevertheless, the power and precision of MR is driven by the heritability of metabolites, due to its dependence on genetic instruments, and some truly causal metabolites may have low power. Second, because the MR analyses use genetic instruments as instrumental variables, they capture estimates based on lifelong genetic exposures without consideration for acute or temporary effects. For example, MR does not account for the effects of initiating statin therapy in older ages. Third, MR infers causality based on observational data, therefore provides causal estimates without definitively establishing causality. Fourth, the UK Biobank is predominantly composed of individuals from European ancestry, so the generalizability of our findings to other ancestry groups may be limited.

Taken together, we found that metabolomic profiles identified via MR are shared across CAD and PAD, but not VTE, suggesting similarities in plaque formation over thrombus formation with respect to blood-based metabolomics. In addition, this suggests that the similarities in plaque formation (e.g., CAD and PAD) outweigh similarities in location (e.g., coronary arteries for CAD, and peripheral beds for PAD and VTE). Atherosclerotic mechanisms, such as VLDL pathways, may be implicated in plaque formation, but not thrombus formation.

## Data Availability

Data can be located upon approval by the UK Biobank, CARDIoGRAMC4D, Million Veterans Project, as well as at the IEU GWAS database (https://gwas.mrcieu.ac.uk/).

## Acknowledgements

Dr. Gilliland was partially supported by the National Heart, Lung, and Blood Institute T32 grant 5T32HL125232. Dr. Nakao is supported by the Japan Society for the Promotion of Science Overseas Fellowship. Dr. Peloso is support by grants from the National Heart Lung and Blood Institute (R01HL142711, R01HL127564). Dr. Natarajan is supported by grants from the National Heart Lung and Blood Institute (R01HL142711, R01HL127564, R01HL148050, R01HL151283, R01HL148565, R01HL135242, R01HL151152), National Institute of Diabetes and Digestive and Kidney Diseases (R01DK125782), Fondation Leducq (TNE-18CVD04), and Massachusetts General Hospital (Paul and Phyllis Fireman Endowed Chair in Vascular Medicine).

## Ethics Declarations

P.N. reports research grants from Allelica, Apple, Amgen, Boston Scientific, Genentech / Roche, and Novartis, personal fees from Allelica, Apple, AstraZeneca, Blackstone Life Sciences, Foresite Labs, Genentech / Roche, GV, HeartFlow, Magnet Biomedicine, and Novartis, scientific advisory board membership of Esperion Therapeutics, Preciseli, and TenSixteen Bio, scientific co-founder of TenSixteen Bio, equity in Preciseli and TenSixteen Bio, and spousal employment at Vertex Pharmaceuticals, all unrelated to the present work.

## Supplement

**Supplementary Figure 1.**
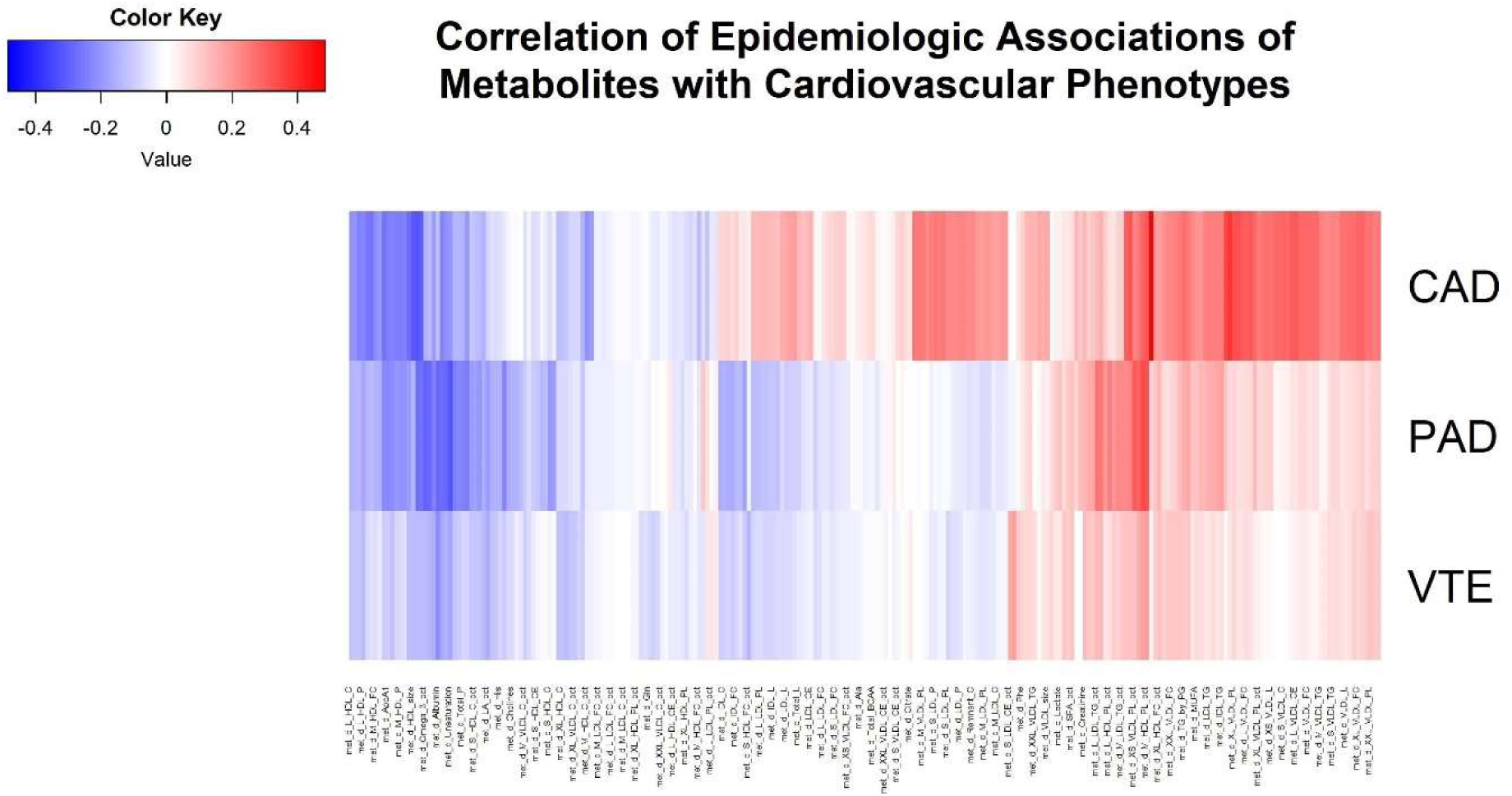
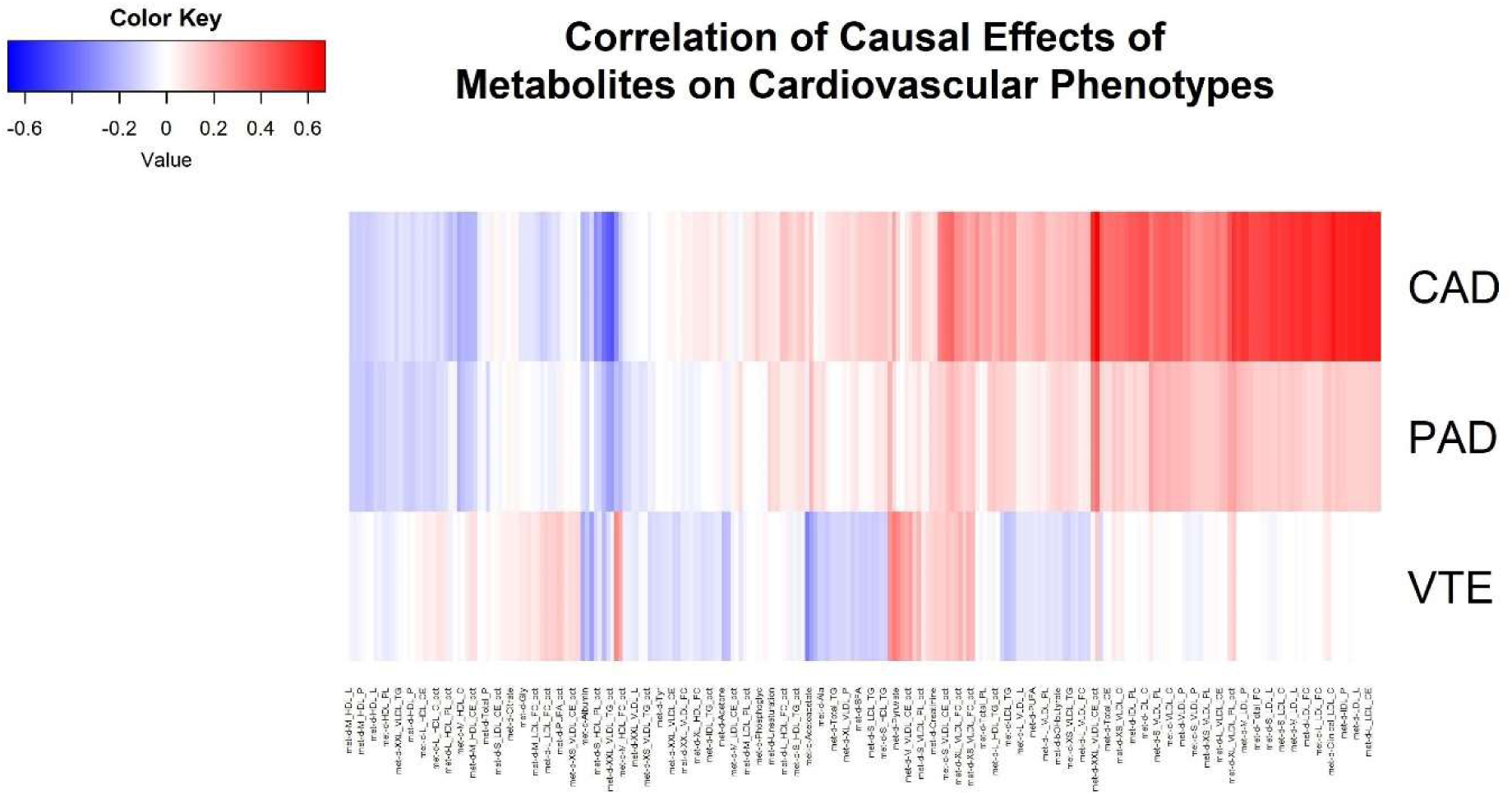
Heatmap summarizing correlation of (A) epidemiologic associations and (B) causal effects of metabolites on cardiovascular phenotypes. (A) Epidemiologic associations were estimated using logistic regression on 95,402 individuals in the UK Biobank. Epidemiologic metabolomic profiles were most similar between PAD and VTE (R^2^ = 0.75, P = 8.43×10^-775^), followed by CAD and PAD (R^2^ = 0.50, P = 5.71×10^-395^) and CAD and VTE (R^2^ = 0.46, P = 2.14×10^-345^). (B) Causal effects were estimated using Mendelian randomization using summary statistics from the UK Biobank and CARDIoGRAMplusC4D 2015 or Million Veterans Program (MVP). Causal metabolomic profiles were most similar between CAD and PAD (R^2^ = 0.43, P = 4.67×10^-156^), followed by PAD and VTE (R^2^ = 0.06, P = 3.55×10^-17^) and CAD and VTE (R^2^ = 0.03, P = 9.38×10^-11^).

**Supplementary Figure 2.**
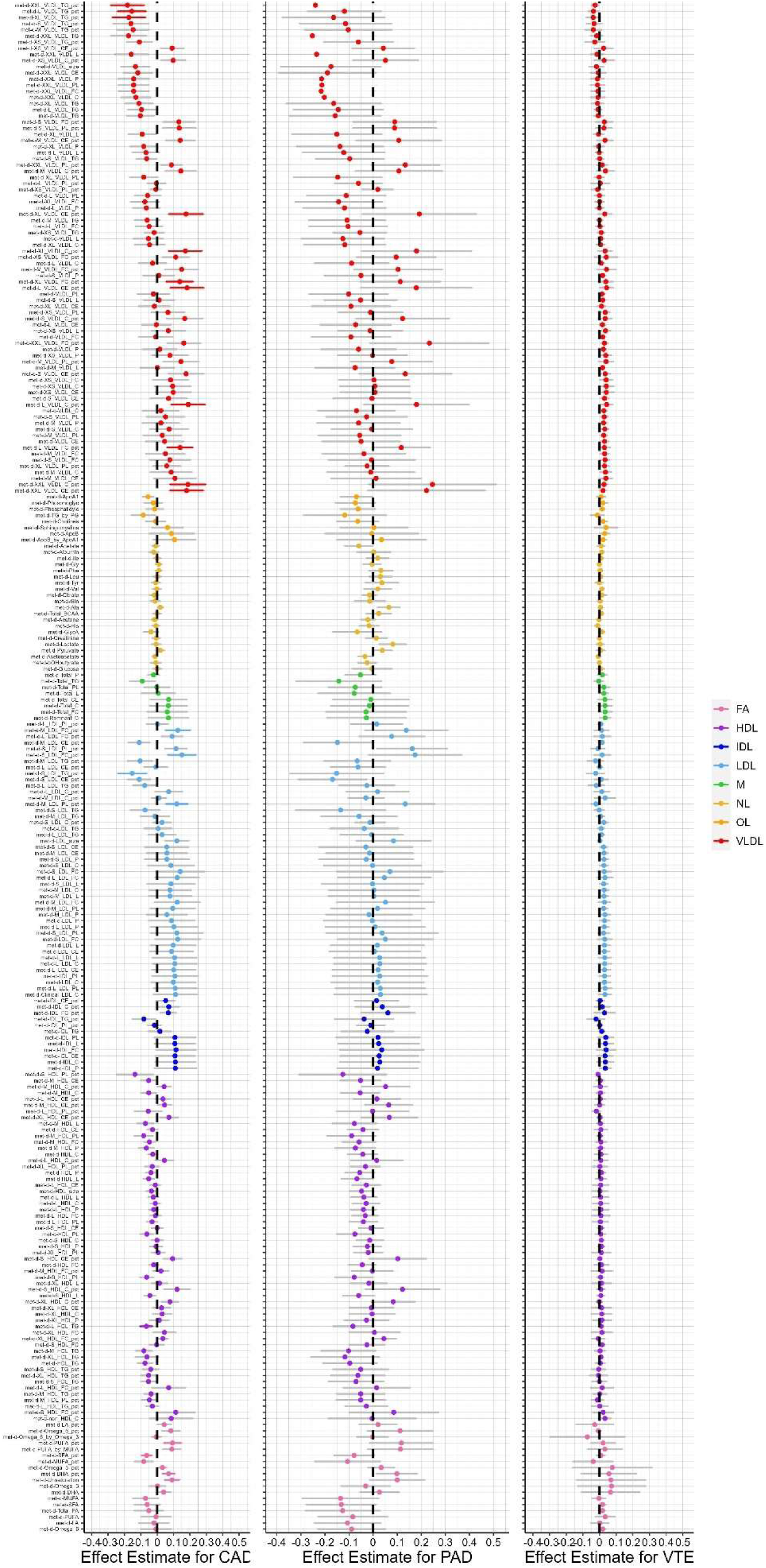
Forest plots of estimated causal effects of cardiovascular outcomes on metabolites estimated using bidirectional Mendelian randomization. Bidirectional, two-sample inverse variance weighted Mendelian randomization using summary statistics from the UK Biobank and CARDIoGRAMplusC4D 2015 or Million Veterans Program (MVP) was performed to estimate causal effects of coronary artery disease (CAD), peripheral artery disease (PAD), and venous thromboembolism (VTE) on metabolites. Bars are effect estimates from Mendelian randomization. Error bars represent 95% confidence intervals. Transparent bars represent associations that were not significant based on a multiple testing correction of P < 0.05 / 41. Only 20 metabolites were causally affected by a cardiovascular outcome, CAD.

**Supplementary Figure 3.**
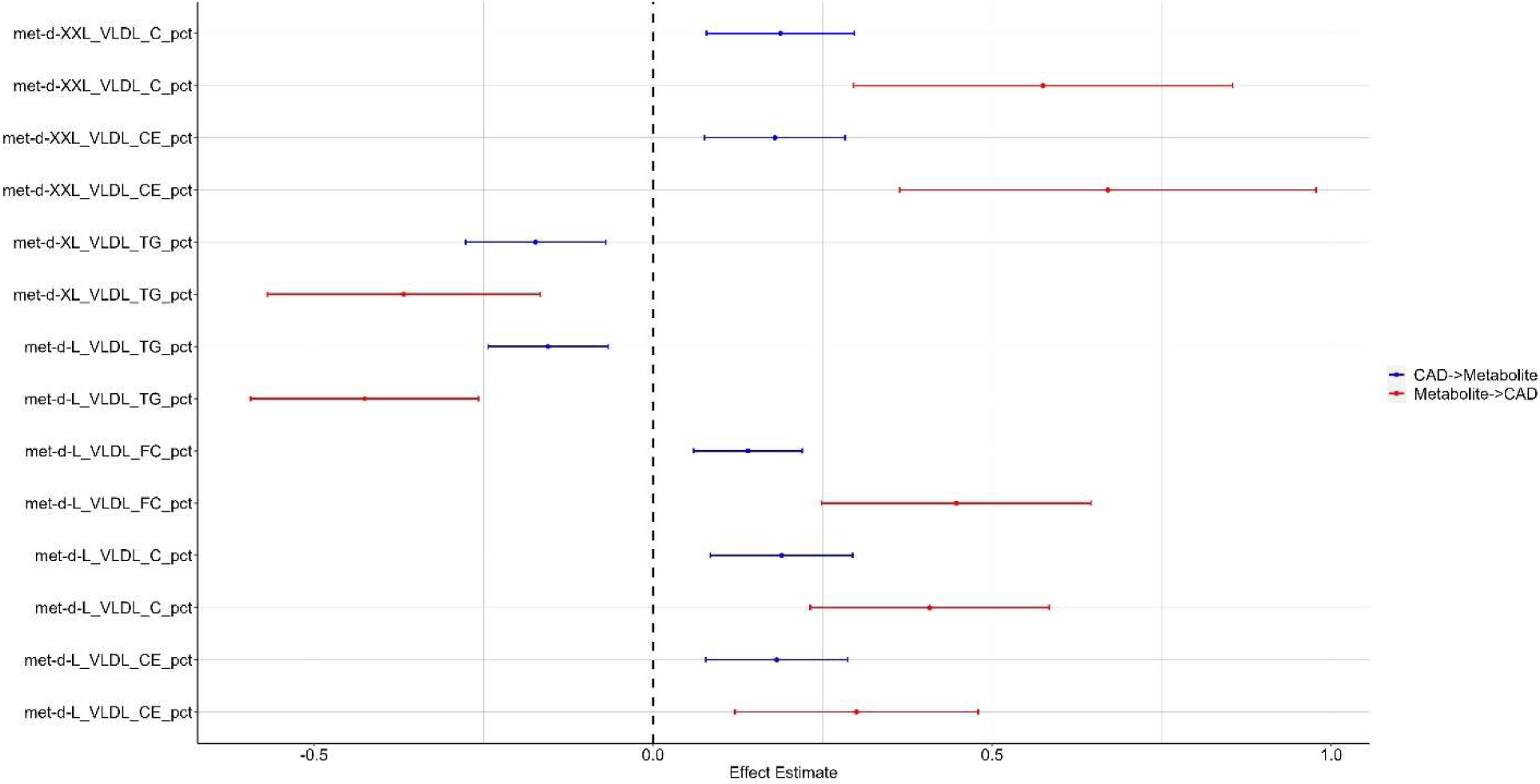
Forest plots of estimated causal effects of metabolites on coronary artery disease and vice versa estimated using bidirectional Mendelian randomization. Bidirectional, two-sample inverse variance weighted Mendelian randomization using summary statistics from the UK Biobank and CARDIoGRAMplusC4D 2015 was performed to estimate causal effects of metabolites on coronary artery disease (CAD) and vice versa. Bars are effect estimates from Mendelian randomization. Error bars represent 95% confidence intervals. Of 7 metabolites with bidirectional causal effects, 2 metabolites decreased risk of CAD and also had levels affected by CAD downstream (e.g., having CAD decreased levels of the metabolite) and 5 metabolites increased risk of CAD and also had levels affected by CAD downstream (e.g., having CAD increased levels of the metabolite.

**Supplementary Table 1.** Phenotype definitions for coronary artery disease (CAD) in CARDIoGRAMplusC4D 2015, peripheral artery disease (PAD) in Million Veterans Program (MVP), and venous thromboembolism (VTE) in MVP.

**Supplementary Table 2.** Summary of instrumental variables for metabolites in the UK Biobank.

**Supplementary Table 3** Table of association analysis estimates between metabolites and coronary artery disease (CAD), peripheral artery disease (PAD), and venous thromboembolism (VTE). Associations were estimated using logistic regression on 95,402 individuals in the UK Biobank.

**Supplementary Table 4.** Table of bidirectional, two-sample Mendelian randomization using the MR Egger, weighted median, inverse variance weighted, simple mode, and weighted mode method estimates using metabolite summary statistics from the UK Biobank as exposures and coronary artery disease (CAD) summary statistics from CARDIoGRAMplusC4D 2015 as the outcome.

**Supplementary Table 5.** Table of bidirectional, two-sample Mendelian randomization using the MR Egger, weighted median, inverse variance weighted, simple mode, and weighted mode method estimates using metabolite summary statistics from the UK Biobank as outcomes and coronary artery disease (CAD) summary statistics from CARDIoGRAMplusC4D 2015 as the exposure.

**Supplementary Table 6.** Table of bidirectional, two-sample Mendelian randomization using the MR Egger, weighted median, inverse variance weighted, simple mode, and weighted mode method estimates using metabolite summary statistics from the UK Biobank as exposures and peripheral artery disease (PAD) summary statistics from Million Veterans Program (MVP) as the outcome.

**Supplementary Table 7.** Table of bidirectional, two-sample Mendelian randomization using the MR Egger, weighted median, inverse variance weighted, simple mode, and weighted mode method estimates using metabolite summary statistics from the UK Biobank as outcomes and peripheral artery disease (PAD) summary statistics from Million Veterans Program (MVP) as the exposure.

**Supplementary Table 8.** Table of bidirectional, two-sample Mendelian randomization using the MR Egger, weighted median, inverse variance weighted, simple mode, and weighted mode method estimates using metabolite summary statistics from the UK Biobank as exposures and venous thromboembolism (VTE) summary statistics from Million Veterans Program (MVP) as the outcome.

**Supplementary Table 9.** Table of bidirectional, two-sample Mendelian randomization using the MR Egger, weighted median, inverse variance weighted, simple mode, and weighted mode method estimates using metabolite summary statistics from the UK Biobank as outcomes and venous thromboembolism (VTE) summary statistics from Million Veterans Program (MVP) as the exposure.

**Supplementary Table 10.** Table of sensitivity analysis results assessing heterogeneity for two-sample Mendelian randomization using metabolites summary statistics from the UK Biobank as exposures and coronary artery disease (CAD), peripheral artery disease (PAD), and venous thromboembolism (VTE) summary statistics from CARDIoGRAMplusC4D 2015 and Million Veterans Program (MVP) as outcomes.

**Supplementary Table 11.** Table of sensitivity analysis results assessing horizontal pleiotropy for two-sample Mendelian randomization using metabolites summary statistics from the UK Biobank as exposures and coronary artery disease (CAD), peripheral artery disease (PAD), and venous thromboembolism (VTE) summary statistics from CARDIoGRAMplusC4D 2015 and Million Veterans Program (MVP) as outcomes.

**Supplementary Table 12.** Table of multivariable Mendelian randomization using 87 sets of metabolite summary statistics from the UK Biobank as exposures and coronary artery disease (CAD) summary statistics from CARDIoGRAMplusC4D 2015 as the outcome. Metabolite sets were determined by selecting the metabolites with significant causal effects on CAD in univariable Mendelian randomization such that genetic instruments included pool of all genetic variants associated with the 87 metabolites.

**Supplementary Table 13.** Table of multivariable Mendelian randomization using 36 sets of metabolite summary statistics from the UK Biobank as exposures and peripheral artery disease (PAD) summary statistics from Million Veterans Program (MVP) as the outcome. Metabolite sets were determined by selecting the metabolites with significant causal effects on PAD in univariable Mendelian randomization such that genetic instruments included pool of all genetic variants associated with the 36 metabolites.

**Supplementary Table 14.** Table of multivariable Mendelian randomization using 2 sets of metabolite summary statistics from the UK Biobank as exposures and venous thromboembolism (VTE) summary statistics from Million Veterans Program (MVP) as the outcome. Metabolite sets were determined by selecting the metabolites with significant causal effects on VTE in univariable Mendelian randomization such that genetic instruments included pool of all genetic variants associated with the 2 metabolites.

## References

1. Mensah GA, Roth GA, Fuster V. The Global Burden of Cardiovascular Diseases and Risk Factors: 2020 and Beyond. J Am Coll Cardiol. Nov 19 2019;74(20):2529–2532. doi:10.1016/j.jacc.2019.10.009

2. Klarin D, Lynch J, Aragam K, et al. Genome-wide association study of peripheral artery disease in the Million Veteran Program. Nat Med. Aug 2019;25(8):1274–1279. doi:10.1038/s41591-019-0492-5

3. Levin MG, Zuber V, Walker VM, et al. Prioritizing the Role of Major Lipoproteins and Subfractions as Risk Factors for Peripheral Artery Disease. Circulation. Aug 03 2021;144(5):353–364. doi:10.1161/CIRCULATIONAHA.121.053797

4. Identification of genetic correlates of coronary artery disease in diverse ancestral populations. Nat Med. Aug 2022;28(8):1548–1549. doi:10.1038/s41591-022-01915-y

5. Aragam KG, Jiang T, Goel A, et al. Discovery and systematic characterization of risk variants and genes for coronary artery disease in over a million participants. Nat Genet. Dec 2022;54(12):1803–1815. doi:10.1038/s41588-022-01233-6

6. Klarin D, Busenkell E, Judy R, et al. Genome-wide association analysis of venous thromboembolism identifies new risk loci and genetic overlap with arterial vascular disease. Nat Genet. Nov 2019;51(11):1574–1579. doi:10.1038/s41588-019-0519-3

7. Poredos P, Jug B. The prevalence of peripheral arterial disease in high risk subjects and coronary or cerebrovascular patients. Angiology. 2007;58(3):309–15. doi:10.1177/0003319707302494

8. Iliou A, Mikros E, Karaman I, et al. Metabolic phenotyping and cardiovascular disease: an overview of evidence from epidemiological settings. Heart. Jul 2021;107(14):1123–1129. doi:10.1136/heartjnl-2019-315615

9. Julkunen H, Cichońska A, Tiainen M, et al. Atlas of plasma NMR biomarkers for health and disease in 118,461 individuals from the UK Biobank. Nat Commun. Feb 03 2023;14(1):604. doi:10.1038/s41467-023-36231-7

10. Yu Z, Zekavat SM, Haidermota S, et al. Genome-wide pleiotropy analysis of coronary artery disease and pneumonia identifies shared immune pathways. Sci Adv. Apr 22 2022;8(16):eabl4602. doi:10.1126/sciadv.abl4602

11. Smith GD, Ebrahim S. ’Mendelian randomization’: can genetic epidemiology contribute to understanding environmental determinants of disease? Int J Epidemiol. Feb 2003;32(1):1–22. doi:10.1093/ije/dyg070

12. Fang S, Holmes MV, Gaunt TR, Davey Smith G, Richardson TG. Constructing an atlas of associations between polygenic scores from across the human phenome and circulating metabolic biomarkers. Elife. Oct 11 2022;11doi:10.7554/eLife.73951

13. Julkunen H, Cichońska A, Slagboom PE, Würtz P, Initiative NHUB. Metabolic biomarker profiling for identification of susceptibility to severe pneumonia and COVID-19 in the general population. Elife. May 04 2021;10doi:10.7554/eLife.63033

14. Sudlow C, Gallacher J, Allen N, et al. UK Biobank: An Open Access Resource for Identifying the Causes of a Wide Range of Complex Diseases of Middle and Old Age. PLoS medicine. Mar 2015;12(3):e1001779. doi:10.1371/journal.pmed.1001779

15. Nikpay M, Goel A, Won HH, et al. A comprehensive 1,000 Genomes-based genome-wide association meta-analysis of coronary artery disease. Nat Genet. Oct 2015;47(10):1121–1130. doi:10.1038/ng.3396

16. Bulik-Sullivan B, Finucane HK, Anttila V, et al. An atlas of genetic correlations across human diseases and traits. Nat Genet. Nov 2015;47(11):1236–41. doi:10.1038/ng.3406

17. Scheinin I, Kalimeri M, Jagerroos V, et al. NMR Data Analysis Tutorial. 2020.

18. Hartwig FP, Davies NM, Hemani G, Davey Smith G. Two-sample Mendelian randomization: avoiding the downsides of a powerful, widely applicable but potentially fallible technique. Int J Epidemiol. 12 01 2016;45(6):1717–1726. doi:10.1093/ije/dyx028

19. Borges MC, Haycock PC, Zheng J, et al. Role of circulating polyunsaturated fatty acids on cardiovascular diseases risk: analysis using Mendelian randomization and fatty acid genetic association data from over 114,000 UK Biobank participants. BMC Med. Jun 13 2022;20(1):210. doi:10.1186/s12916-022-02399-w

20. Hemani G, Zheng J, Elsworth B, et al. The MR-Base platform supports systematic causal inference across the human phenome. Elife. 05 30 2018;7doi:10.7554/eLife.34408

21. Hemani G, Tilling K, Davey Smith G. Orienting the causal relationship between imprecisely measured traits using GWAS summary data. PLoS Genet. Nov 2017;13(11):e1007081. doi:10.1371/journal.pgen.1007081

22. Burgess S, Davey Smith G, Davies NM, et al. Guidelines for performing Mendelian randomization investigations. Wellcome Open Res. 2019;4:186. doi:10.12688/wellcomeopenres.15555.2

23. Burgess S, Butterworth A, Thompson SG. Mendelian randomization analysis with multiple genetic variants using summarized data. Genet Epidemiol. Nov 2013;37(7):658–65. doi:10.1002/gepi.21758

24. Burgess S, Bowden J, Fall T, Ingelsson E, Thompson SG. Sensitivity Analyses for Robust Causal Inference from Mendelian Randomization Analyses with Multiple Genetic Variants. Epidemiology. Jan 2017;28(1):30–42. doi:10.1097/EDE.0000000000000559

25. Burgess S, Thompson SG. Multivariable Mendelian randomization: the use of pleiotropic genetic variants to estimate causal effects. Am J Epidemiol. Feb 15 2015;181(4):251–60. doi:10.1093/aje/kwu283

26. Jia A, Zeng W, Yu L, Zeng H, Lu Z, Song Y. Very low-density lipoprotein cholesterol is associated with extent and severity of coronary artery disease in patients with type 2 diabetes mellitus. SAGE Open Med. 2019;7:2050312119871786. doi:10.1177/2050312119871786

27. Aday AW, Everett BM. Dyslipidemia Profiles in Patients with Peripheral Artery Disease. Curr Cardiol Rep. Apr 22 2019;21(6):42. doi:10.1007/s11886-019-1129-5

28. National Cholesterol Education Program (NCEP) Expert Panel on Detection Ea, and Treatment of High Blood Cholesterol in Adults (Adult Treatment Panel III). Third Report of the National Cholesterol Education Program (NCEP) Expert Panel on Detection, Evaluation, and Treatment of High Blood Cholesterol in Adults (Adult Treatment Panel III) final report. Circulation. Dec 17 2002;106(25):3143–421.

29. Nordestgaard BG. Triglyceride-Rich Lipoproteins and Atherosclerotic Cardiovascular Disease: New Insights From Epidemiology, Genetics, and Biology. Circ Res. Feb 19 2016;118(4):547–63. doi:10.1161/CIRCRESAHA.115.306249

30. Talayero BG, Sacks FM. The role of triglycerides in atherosclerosis. Curr Cardiol Rep. Dec 2011;13(6):544–52. doi:10.1007/s11886-011-0220-3

31. Franczyk B, Gluba-Brzózka A, Ławiński J, Rysz-Górzyńska M, Rysz J. Metabolomic Profile in Venous Thromboembolism (VTE). Metabolites. Jul 29 2021;11(8)doi:10.3390/metabo11080495

32. Lin L, Luo P, Yang M, Wang J, Hou W, Xu P. A bidirectional Mendelian randomized study of classical blood lipids and venous thrombosis. Sci Rep. Mar 08 2023;13(1):3904. doi:10.1038/s41598-023-31067-z

33. Tomas L, Edsfeldt A, Mollet IG, et al. Altered metabolism distinguishes high-risk from stable carotid atherosclerotic plaques. Eur Heart J. Jun 21 2018;39(24):2301–2310. doi:10.1093/eurheartj/ehy124

